# A geotemporal survey of hospital bed saturation across England during the first wave of the COVID-19 Pandemic

**DOI:** 10.1101/2020.06.24.20139048

**Authors:** Bilal A. Mateen, Harrison Wilde, John M. Dennis, Andrew Duncan, Nicholas J. Thomas, Andrew P. McGovern, Spiros Denaxas, Matt J Keeling, Sebastian J. Vollmer

**Author notes:** **<u>Author for Correspondence</u>** Dr. Bilal A. Mateen, Social Science and Systems in Health Unit, Warwick Medical School, University of Warwick, Coventry, CV4 7AL, UK.

## Abstract

**Background:** Non-pharmacological interventions were introduced based on modelling studies which suggested that the English National Health Service (NHS) would be overwhelmed by the COVID-19 pandemic. In this study, we describe the pattern of bed occupancy across England during the first wave of the pandemic, January 31st to June 5th 2020.

**Methods:** Bed availability and occupancy data was extracted from daily reports submitted by all English secondary care providers, between 27-Mar and 5-June. Two thresholds for ‘safe occupancy’ were utilized (85% as per Royal College of Emergency Medicine and 92% as per NHS Improvement).

**Findings:** At peak availability, there were 2711 additional beds compatible with mechanical ventilation across England, reflecting a 53% increase in capacity, and occupancy never exceeded 62%. A consequence of the repurposing of beds meant that at the trough, there were 8·7% (8,508) fewer general and acute (G&A) beds across England, but occupancy never exceeded 72%. The closest to (surge) capacity that any trust in England reached was 99·8% for general and acute beds. For beds compatible with mechanical ventilation there were 326 trust-days (3·7%) spent above 85% of surge capacity, and 154 trust-days (1·8%) spent above 92%. 23 trusts spent a cumulative 81 days at 100% saturation of their surge ventilator bed capacity (median number of days per trust = 1 [range: 1 to 17]). However, only 3 STPs (aggregates of geographically co-located trusts) reached 100% saturation of their mechanical ventilation beds.

**Interpretation:** Throughout the first wave of the pandemic, an adequate supply of all bed-types existed at a national level. Due to an unequal distribution of bed utilization, many trusts spent a significant period operating above ‘safe-occupancy’ thresholds, despite substantial capacity in geographically co-located trusts; a key operational issue to address in preparing for a potential second wave.

**Funding:** This study received no funding.

**Research In Context:** *Evidence Before This Study:* We identified information sources describing COVID-19 related bed and mechanical ventilator demand modelling, as well as bed occupancy during the first wave of the pandemic by performing regular searches of MedRxiv, PubMed and Google, using the terms ‘COVID-19’, ‘mechanical ventilators’, ‘bed occupancy’, ‘England’, ‘UK’, ‘demand’, and ‘non-pharmacological interventions (NPIs)’, until June 20th, 2020. Two UK-specific studies were found that modelled the demand for mechanical ventilators, one of which incorporated sensitivity analysis based on the introduction of NPIs and found that their effects might prevent the healthcare system being overwhelmed. Separately, several news reports were found pertaining to a single hospital that reached ventilator capacity in England during the first wave of the pandemic, however, no single authoritative source was identified detailing impact across all hospital sites in England.

*Added Value of This Study:* This national study of hospital-level bed occupancy in England provides unique and timely insight into bed-specific resource utilization during the first wave of the COVID-19 pandemic, nationally, and by specific (geographically defined) health footprints. We found evidence of an unequal distribution of resource utilization across England. Although occupancy of beds compatible with mechanical ventilation never exceeded 62% at the national level, 52 (30%) hospitals across England reached 100% saturation at some point during the first wave of the pandemic. Close examination of the geospatial data revealed that in the vast majority of circumstances there was relief capacity in geographically co-located hospitals. Over the first wave it was theoretically possible to markedly reduce (by 95.1%) the number of hospitals at 100% saturation of their mechanical ventilator bed capacity by redistributing patients to nearby hospitals.

*Implications Of All The Available Evidence:* Now-casting using routinely collected administrative data presents a robust approach to rapidly evaluate the effectiveness of national policies introduced to prevent a healthcare system being overwhelmed in the context of a pandemic illness. Early investment in operational field hospital and an independent sector network may yield more overtly positive results in the winter, when G&A occupancy-levels regularly exceed 92% in England, however, during the first wave of the pandemic they were under-utilized. Moreover, in the context of the non-pharmacological interventions utilized during the first wave of COVID-19, demand for beds and mechanical ventilators was much lower than initially predicted, but despite this many trust spent a significant period of time operating above ‘safe-occupancy’ thresholds. This finding demonstrates that it is vital that future demand (prediction) models reflect the nuances of local variation within a healthcare system. Failure to incorporate such geographical variation can misrepresent the likelihood of surpassing availability thresholds by averaging out over regions with relatively lower demand, and presents a key operational issue for policymakers to address in preparing for a potential second wave.

## Introduction

The ability of hospitals to cope with large influxes of patients, either due to a pandemic illness or seasonal increases in respiratory disease exacerbations is in part dictated by the availability of beds.^1^ Since 1987, when formal reporting of the number of hospital beds began in the UK, there has been a sustained decline in the number of available beds across the NHS.^2^ In recent years, this issue has garnered more attention due to the annual ‘winter bed crisis’,^3,4^ where the end of the calendar year heralds a surge in emergency admissions often resulting in hospitals operating well above quality and operational performance tipping points, i.e. 85% or 92% total bed occupancy.^5–7^ The saturation of hospital beds is not only problematic through it impact on the ability of the workforce to deliver high-quality care,^8^ but additionally the bottle-necking of the emergency care workflow has been shown to contribute to suboptimal outcomes for patients,^9^ including increased numbers of healthcare-acquired infections,^10^ and increased mortality.^11–13^

These concerns about the NHS’ ability to cope with large influxes of patients took on a new level of significance in early 2020, when the World Health Organization (WHO) formally declared COVID-19 a pandemic illness, due to its virulence, and the magnitude of the disease’s impact globally.^14^ As early reports from China were published, it became apparent that a relatively large proportion of individuals who contracted COVID-19 required admission to hospital,^15^ for example due to: new oxygen requirements, sepsis, acute respiratory distress syndrome (ARDS), and even multi-organ dysfunction (MODS). Forecasts of the potential number of people requiring hospital admission and mechanical ventilation across the UK suggested that the baseline capacity of the NHS would be insufficient.^16^ In an effort to ensure sufficient capacity the British government instituted a series of policies, including facilitating the discharge of individuals who had been delayed due to non-medical reasons in an effort to unlock capacity,^17^ cancelling all non-urgent clinical work, opening large field hospitals (i.e. the Nightingale hospitals),^18^ and increasing mechanical ventilator availability for clinical areas repurposed to manage high care patients.^19^

The UK has started making significant strides towards rolling back its non-pharmacological interventions including. reopening schools, and planning for the discontinuation of shielding for vulnerable people,^20^ signaling an end to the first wave of the pandemic.^21^ Following these changes, there is the potential for a second wave of infections in the coming months. Understanding regional differences in hospital capacity is fundamental to informing the UK’s response to a second wave, as well as for elucidating how to safely wind down repurposed surge capacity such as operating theatres to allow other much needed clinical activity to restart.^22^ However, other than a few isolated news reports of hospitals exceeding their ventilator capacity,^23^ it is unclear how well the NHS as a whole managed to respond to the additional demand for beds over recent months. In this study, we sought to describe the pattern of bed occupancy in hospitals across England during the first wave of the COVID-19 pandemic.

## Methods

### Primary Data Source

Data were accessed from the daily situation reports (‘SitReps’, covering the previous 24 hours) provided to the Scientific Pandemic Influence Group on Modelling (SPI-M) by NHS England on behalf of all secondary care providers. All NHS acute care providers, independent sector care providers, and field hospitals in England submitting information to the daily situation reports were eligible for inclusion.

### Study population

The data is presented in the context of several different units of secondary care provision: hospitals/sites, trusts, sustainability and transformation partnerships (STPs), regions, and the whole of England (i.e. national), where each is an aggregate of the preceding unit (the structure of UK care providers is explained in the supplementary material).

#### Inclusion and Exclusion Criteria

Exclusions were applied at the trust level for NHS-specific care providers. Exclusion criteria were as follows: acute specialist trusts: women and/or children (n = 4), neurology & ophthalmology (n = 2), heart & lung (n = 3), orthopedic, burns & plastics (n = 4), cancer (n = 3). The remaining care providers were grouped into three categories and analyzed separately: 1) Acute (non-specialist) Trusts with a type 1 (i.e. 24 hours/day, consultant-led) accident and emergency department (n = 125); 2) Nightingale (Field) Hospitals (n = 7), and; 3) independent sector providers (n = 195).

#### Recruitment Period

Data was available from 27th March 2020 (the first available SitRep) to 5th June 2020 inclusive.

### Recorded Information

The data specification comprised resource utilization and capacity-specific information, including the number of beds at each trust, stratified by several factors of interest, including acuity and COVID-19 ascertainment (further defined in supplementary material). Notably, beds were only recorded as being available if they were ‘funded’ (i.e. there was adequate staffing and resources for the bed to be occupied), so as to prevent counting of beds that could not accommodate a new patient. Bed acuity was organized into: general and acute (G&A), beds compatible with non-mechanical ventilation, and beds compatible with mechanical ventilation. Occupancy is calculated based on the status of each bed at 08:00 each day, and then later separated by the proportion that had a positive COVID-19 test.

Reporting fields changed on the 27th of April 2020, with several additional columns being added, which included specific fields for level 2 (HDU) and level 3 (ITU) beds. The impact of these changes is detailed in the supplementary material. However, one crucial outcome was that it became apparent the definition of critical care beds utilized prior to 27th April 2020 was not consistent with prior reporting practices of only including level 2 (HDU) and level 3 (ITU) beds,^24^ as the newly reported values did not equal the simultaneously reported critical care values. As such, any results pertaining to critical care, HDU and ITU are reported separately in the supplementary material.

NHS England reports trust-level data, whereas we additionally attempted to disaggregate this information into the individual hospitals that the trusts comprise. Not all of the trusts were amenable to disaggregation from the trust-level data into independently reported sites in the available extracts, resulting in a final sample of 173 unique hospital sites, comprising 91·7% of the total number of ventilated beds and 81·4% of the general and acute beds when compared to trust level. The change in data reporting introduced on the 27th of April 2020 also resulted in variation in information capture; for data prior to the 27th of April, the results available reflect 89·6% of all mechanical ventilator beds and 86·9% of general and acute beds, where data from the 27th onwards the results reflect 93·0% of all mechanical ventilator beds but 77·0% of general and acute beds.

### Outcome

The primary outcomes of interest were bed availability, and bed occupancy by patients with and without COVID-19, for each level of secondary care provision, i.e. hospital, trust, sustainability and transformation partnership (STP). Different ‘safe occupancy’ thresholds were used to interpret the results; 85% as per the Royal College of Emergency Medicine, and 92% as per NHS Improvement. We also compared occupancy against baseline bed occupancy (see supplementary material for definitions), and 100% of surge capacity.

### Statistical Analysis

We generated and reported descriptive summaries (e.g. medians, ranges, counts, proportions) of the data. We reported absolute numbers for hospital, trusts and STPs attaining specific occupancy thresholds. In light of the discordant critical care and HDU/ITU values, this analysis was handled and reported separately (see supplementary material). To capture the temporal aspect of the information available, the number of hospital-days, trust-days, and STP-days above hospital baseline capacity and surge capacities of 85%, 92% and 100% is also reported. Full details on the quality control procedures are reported in the supplementary material (SFigures 1 & SFigure 2). Details on aggregation and disaggregation of geographic information are provided in STable 1 & STable 2. Analysis were carried out in R,^25^ ggplot2 package.^26^ Maps were acquired from the UK’s Office for National Statistics Open Geography Portal.^27^

**Table 1:** The Number of Hospital/Trusts/STPs at each Occupancy Threshold for Different Bed Types

| Bed Type | Organizational Unit | Occupancy Threshold |  |  |  |  |  |
| --- | --- | --- | --- | --- | --- | --- | --- |
|  |  | > 85% |  | > 92% |  | = 100% |  |
|  |  | Number (%) | Median number of days at or above threshold (range) | Number (%) | Median number of days at or above threshold (range) | Number (%) | Median number of days at or above threshold (range) |
| General & Acute | Hospital/Site (n = 173) | 56 (32.4%) | 6 (1 to 45] | 19 (11.0%) | 3 (1 to 19) | 1 (0.6%) | 10 days |
|  | Trust (n = 125) | 30 (24.0%) | 5 (1 to 46) | 14 (11.2%) | 3 (1 to 13) | 0 (0.0%) | - |
|  | Sustainability and Transformation Partnership (n = 42) | 2 (4.8%) | 10 (3 to 17) | 2 (4.8%) | 1 (1 to 1) | 0 (0.0%) | - |
| Mechanical Ventilation | Hospital/Site (n = 173) | 91 (52.6%) | 4 (1 to 48) | 72 (41.6%) | 3 (1 to 48) | 52 (30.0%) | 2 (1 to 48) |
|  | Trust (n = 125) | 58 (46.4%) | 3 (1 to 27) | 40 (32.0%) | 2 (1 to 17) | 23 (18.4%) | 1 (1 to 17) |
|  | Sustainability and Transformation Partnership (n = 42) | 10 (23.8%) | 2 (1 to 11) | 5 (10.4%) | 1 (1 to 6) | 3 (7.1%) | 1 (1 to 2) |

**Table 2:**
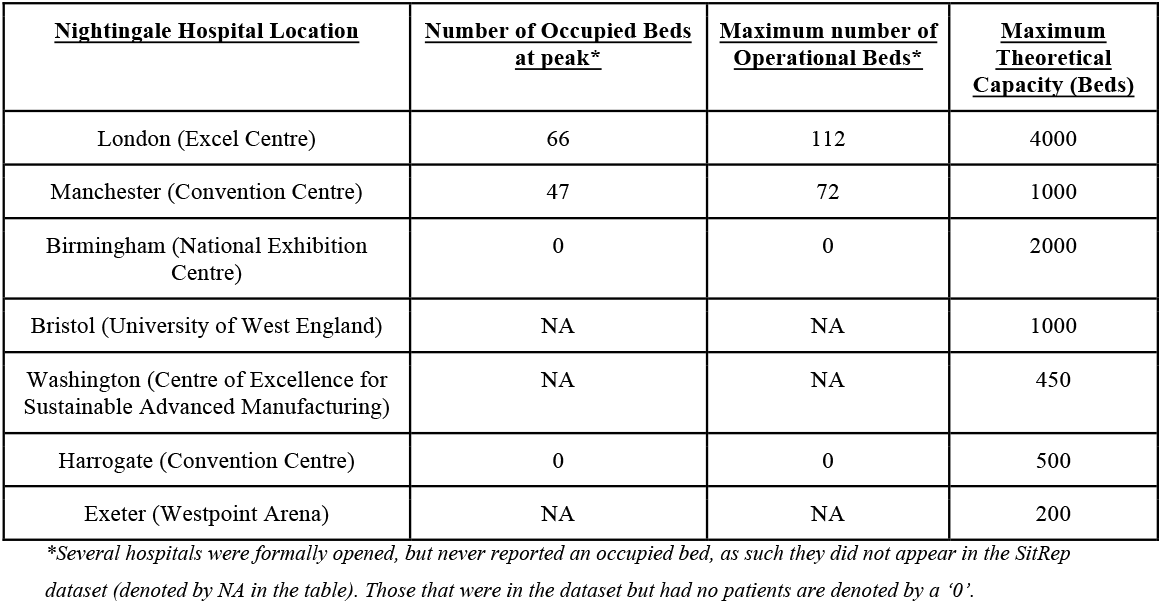
Field (Nightingale) Hospital Occupancy, and Capacity

## Results

### National Mobilization

During the first wave of the pandemic, the NHS repurposed general/acute beds into those suitable for higher acuity patients (i.e. HDU/ITU), and patients requiring mechanical ventilation. Available ventilated bed capacity peaked at an additional 2711 beds, a 53% increase from a baseline of 4123 beds. Ventilated beds occupancy never exceeded 62% of this capacity (Figure 1a), however there were notable regional differences in COVID-19 specific demand (Figure 1b, 1c & SFigure 3). Similar patterns were observed in critical care/HDU and ITU beds (SFigure 4). A consequence of the repurposing of beds for higher acuity patients there was a 8·7% reduction (n= 8,508) of general and acute beds from a baseline of 97,293 beds. There was a large reduction of the number of beds occupied by patients without COVID-19; 53,136 fewer beds were occupied (58·8% reduction) at the nidus compared to average occupancy from January to March 2020. Total bed occupancy never exceeded 72% nationally (Figure 1a).

**Figure 1:**
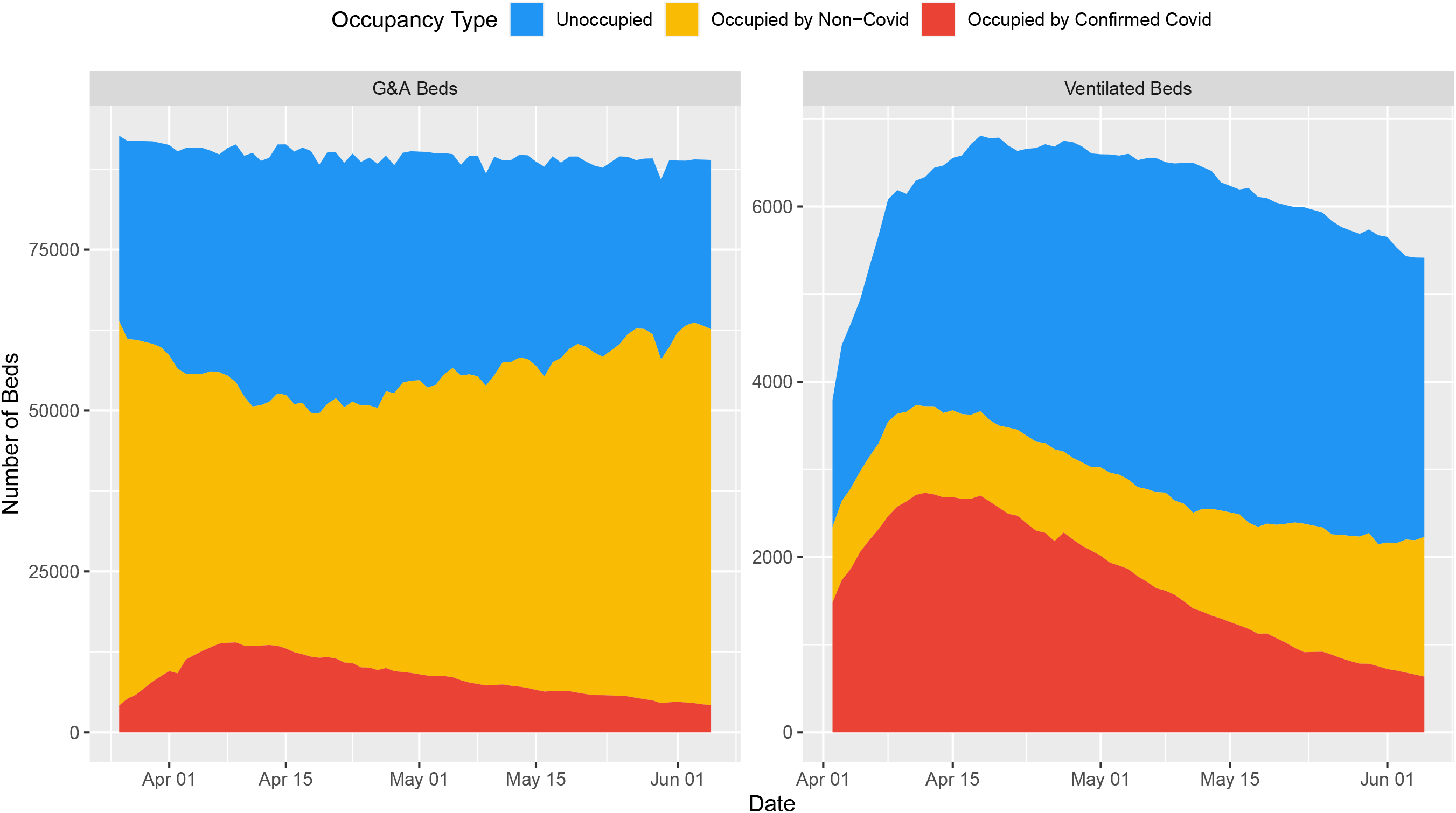
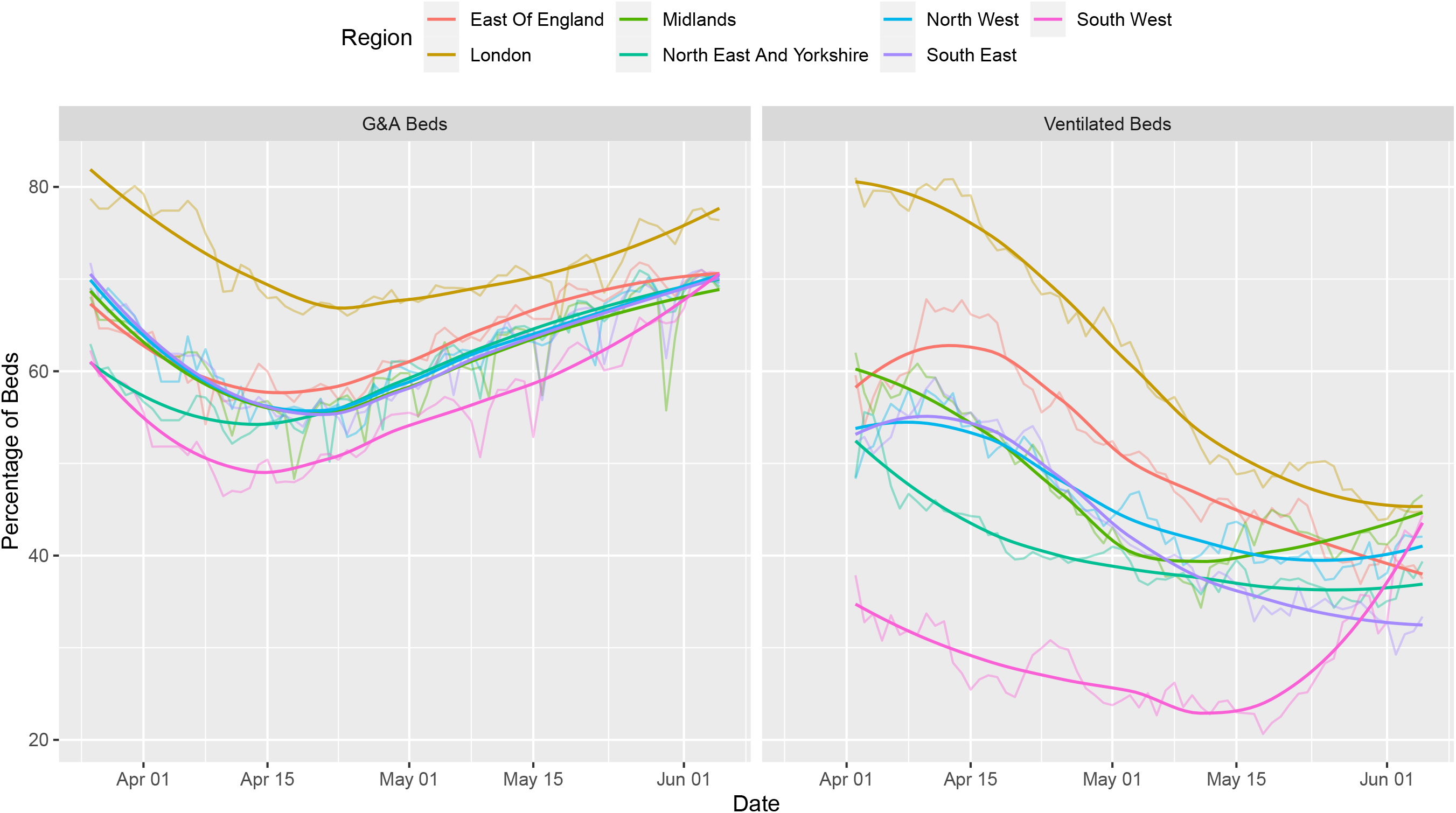
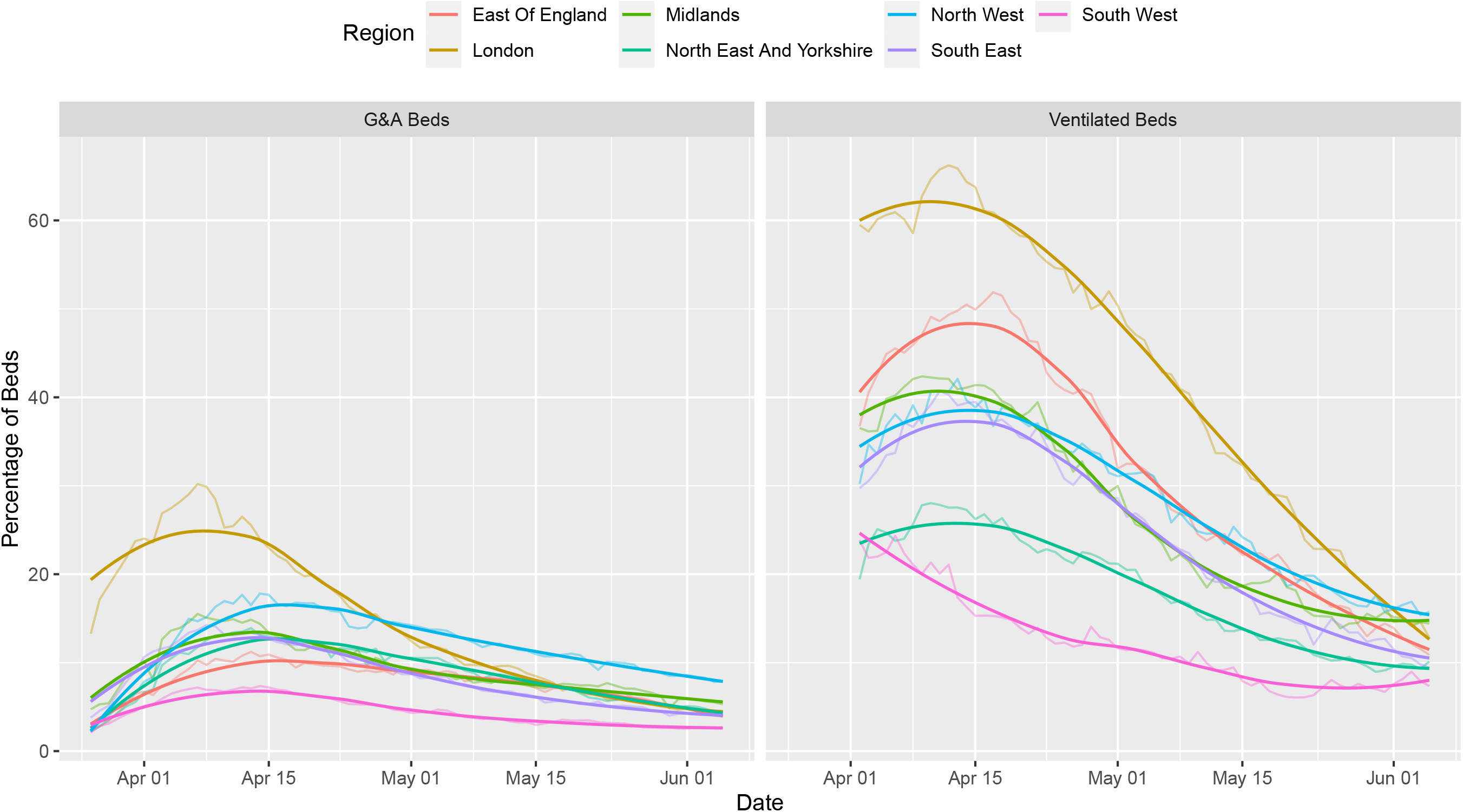
National and Regional Bed Occupancy. Legend: Figure 1A (Top) illustrates total capacity and occupancy status, for both general and acute (G&A), as well as bed compatible with mechanical ventilation, at the national level. Figure 1B (Middle) illustrates total occupancy (COVID-19 positive and negative) in each of the 7 regions across England, for both general and acute (G&A), as well as bed compatible with mechanical ventilation. Figure 1C (Bottom) illustrates COVID-19 specific occupancy in each of the 7 regions across England, for both general and acute (G&A), as well as bed compatible with mechanical ventilation. Note: the highly-saturated solid line represents a smoothened function of the raw data, whereas the less saturated solid line represents the underlying raw values. The former is based on the ggplot loess fit for trend lines, using local poly-regression curve fitting.

### Occupancy relative to Baseline Capacity

Out of the 125 trusts (aggregates of hospitals), 3 trusts (2·4%) at some point during the first wave were operating above their baseline bed availability for general and acute beds (124 trust-days [1·4% of the total 8738 days at risk]; median number of days per trust = 36 days [range: 30 to 58]; Figure 2a). For beds compatible with mechanical ventilation, 87 trusts (69·6%) at some point during the first wave were operating above their baseline bed availability (2456 trust-days [28·1% of total at risk]; median number of days per trust = 24 days [range: 1 to 61]; Figure 2b). Similar results to that of mechanical ventilation compatible beds were seen for critical care / HDU and ITU bed occupancy (see supplementary material, SFigure 5 & SFigure 6).

**Figure 2:**
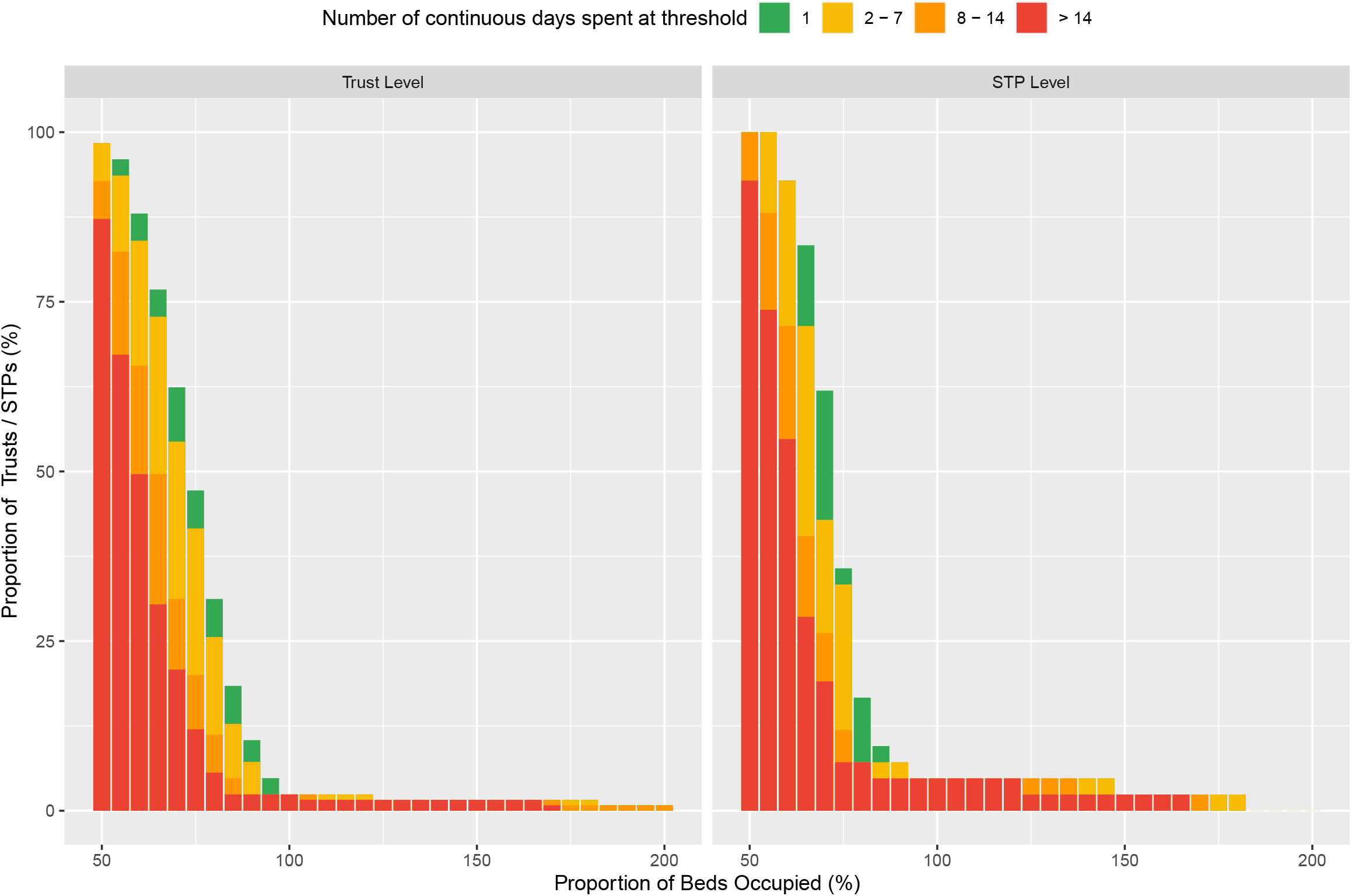
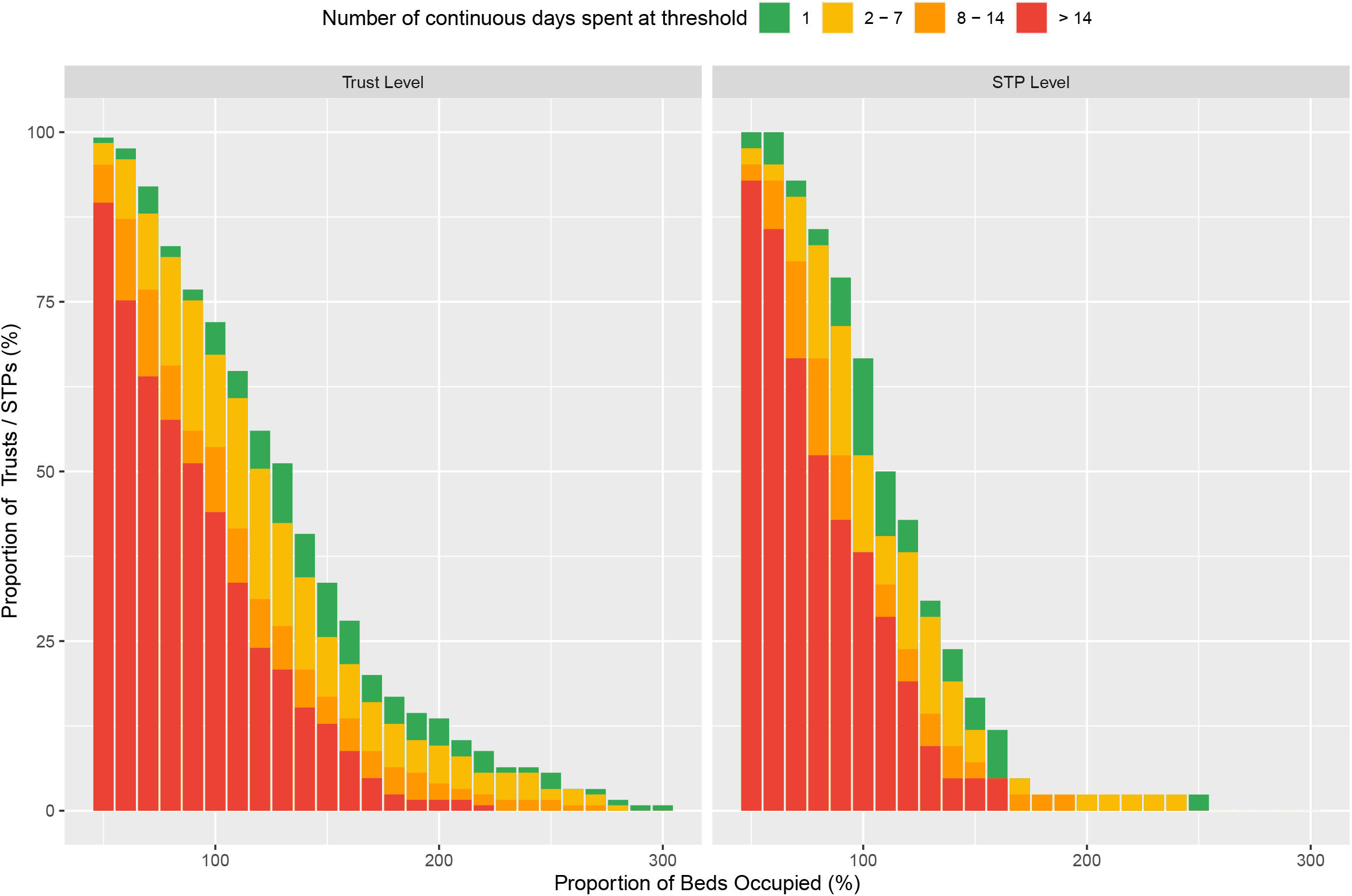
Trust-Level Bed Occupancy (Based on Baseline Capacities) Across England. Legend: Figure 2A (Top) illustrates the proportion of all trusts, and sustainability and transformation partnerships (STPs), at varying general and acute (G&A) bed occupancy thresholds relative to their baseline (mean availability January-March 2020) capacity, across England, from April 1st to June 5th. Figure 2B (Bottom) illustrates the proportion of all trusts, and sustainability and transformation partnerships (STPs), at varying ventilator bed occupancy thresholds relative to their baseline capacity, across England, from April 1st to June 5th. The superimposed colours represent how long the trusts spent at each specific threshold.

### Occupancy relative to Surge Capacity

Table 1 summarizes the number of hospitals, trusts and STPs operating above each of the thresholds for ‘safe occupancy’, and details the duration (i.e. median number of days) that each spent above the designated thresholds.

#### Hospital-level Occupancy

Of the total 11,851 English hospital-days at risk over the study period, 494 hospital-days (4·17% of total days at risk) were at or above 85% of bed (surge) capacity, 110 hospital-days (0·92%) were at or above 92% of bed (surge) capacity, and only 10 were spent at 100% of surge capacity (Figure 3a). Similarly, for beds compatible with mechanical ventilation there were 586 hospital-days (4·94%) spent above 85% of surge capacity, 320 hospital-days (2·70%) were spent above 92%, and 226 hospital-days (1·9%) were spent at 100% occupancy (see Figure 3b). Summaries of the size and geographic locations of hospitals stratified by saturation are in STable 3.

**Figure 3:**
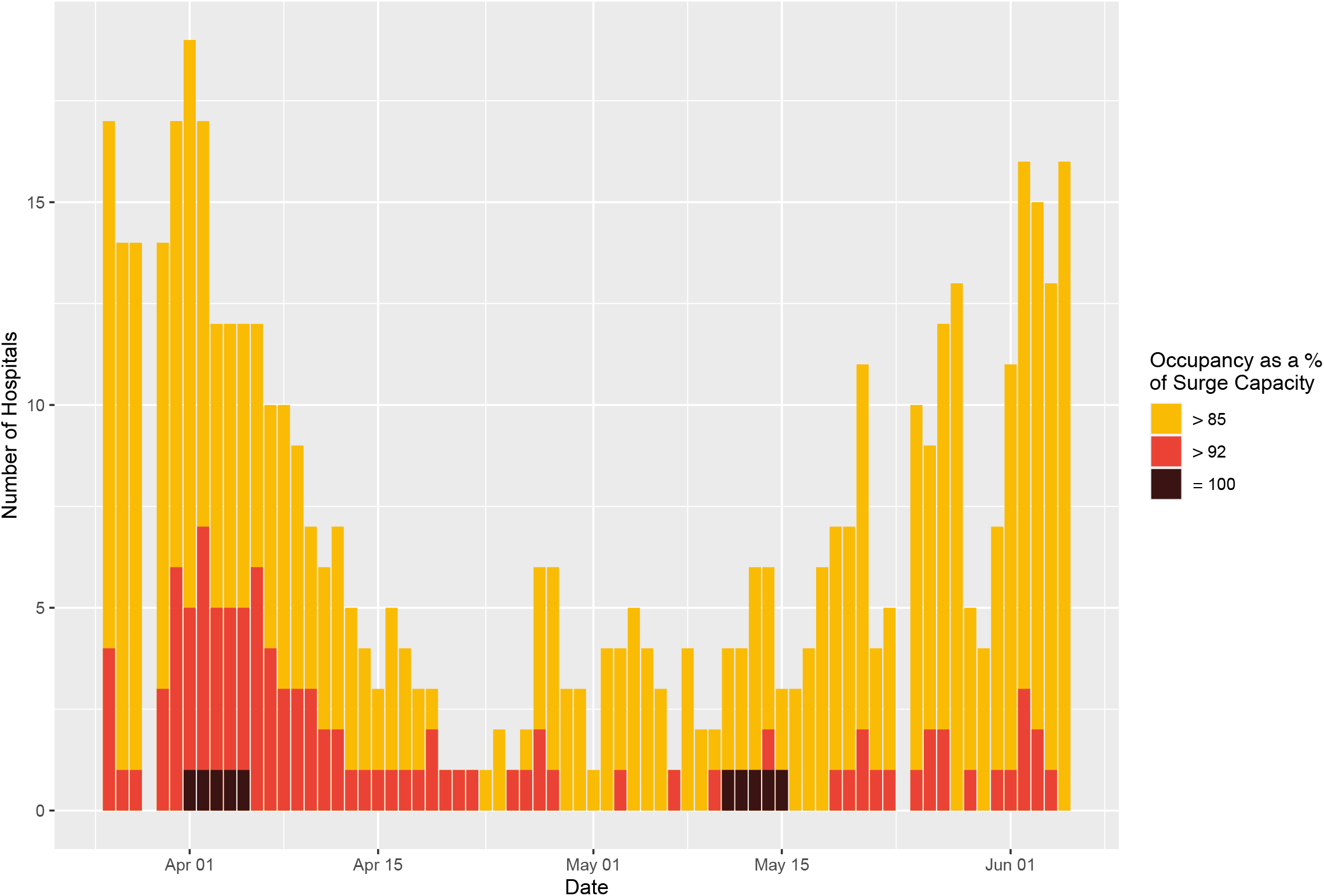
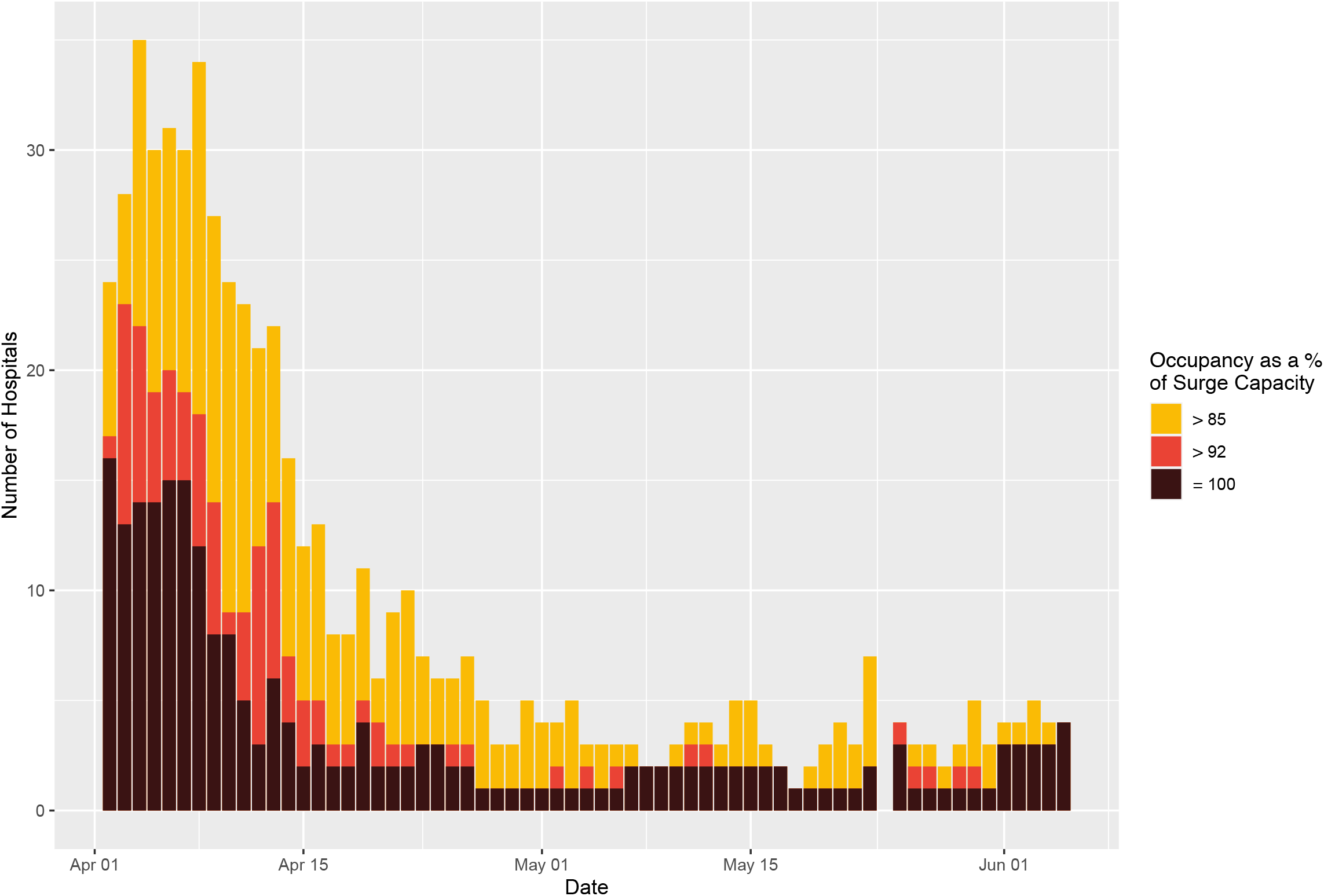
Hospital-level Bed Occupancy (Based on Surge Capacities) Across England. Legend: Figure 3A (Top) illustrates the number of hospitals with general and acute bed occupancy in excess of the thresholds for ‘safe and effective’ functioning, i.e. 85% as defined by the Royal College of Emergency Medicine,^6^ and 92% as defined by NHS Improvement and NHS England (green and yellow, respectively),^7^ across England, from 1st April to June 5th. Figure 3B (Bottom) illustrates the number of hospitals with occupancy of mechanical ventilation beds in excess of the aforementioned thresholds, across England, from April 1st to June 5^th^. Note: all data was missing for the 24th of May.

#### Trust-level Bed Occupancy

Over the study period, there were 287 trust-days (3·3% of total days at risk) where general and acute bed occupancy exceeded 85% occupancy of surge capacity, and 57 trust-days (0·7%) were at or above 92% of bed (surge) capacity. The closest to capacity any trust in England reached was 99.8% for general and acute beds. However, for beds compatible with mechanical ventilation there were 326 trust-days (3·7%) spent above 85% of surge capacity, and 154 trust-days (1·8%) spent above 92%. 23 trusts reached 100% saturation of their mechanical ventilator bed capacity (Figure 4 & SFigure 7).

**Figure 4:**
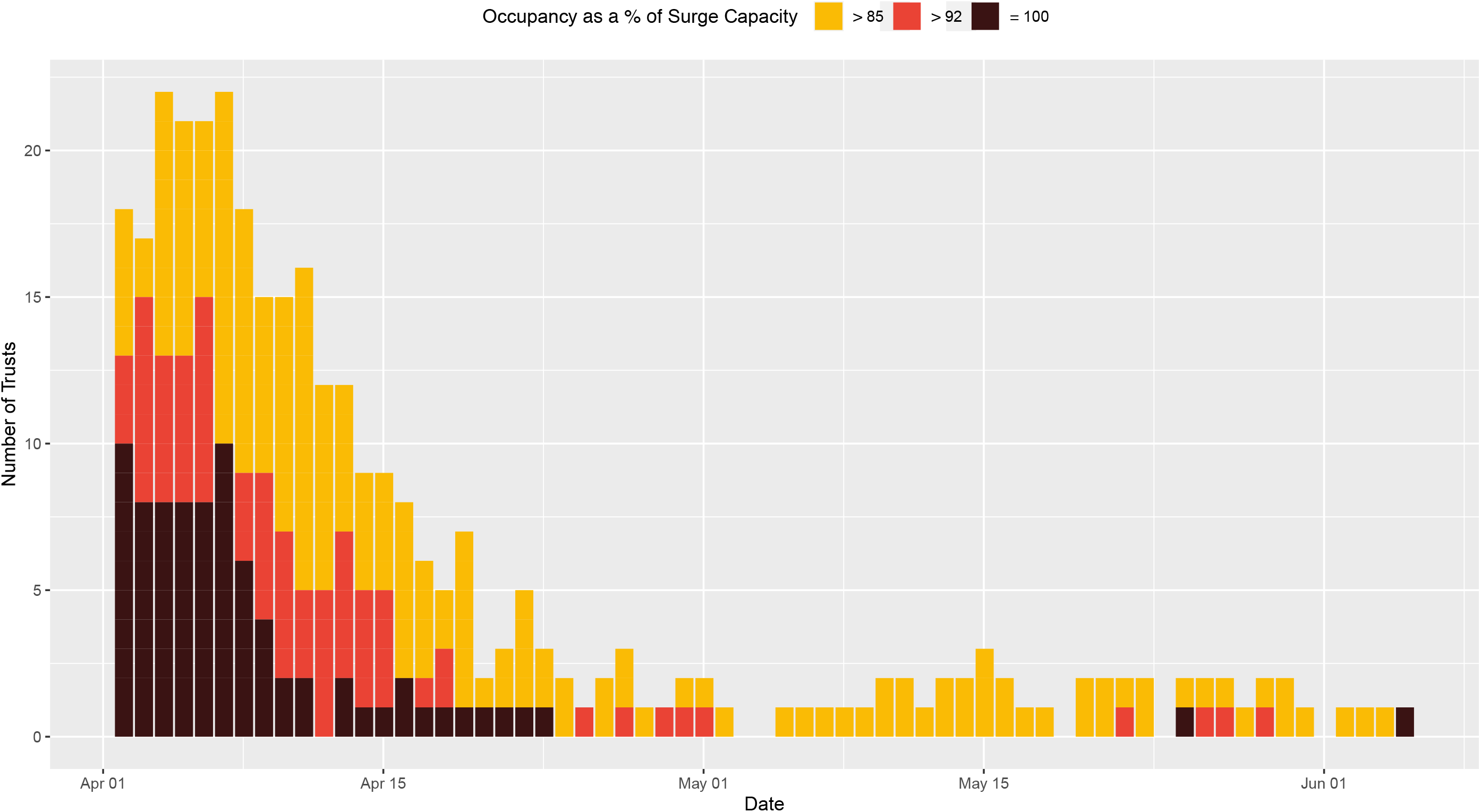
Trust-Level Ventilator Bed Occupancy (Based on Surge Capacities) Across England. Legend: Figure 4 illustrates the number of trusts with occupancy of mechanical ventilation beds in excess of the thresholds for ‘safe and effective’ functioning, i.e. 85% as defined by the Royal College of Emergency Medicine,^6^ and 92% as defined by NHS Improvement and NHS England (yellow and red, respectively),^7^ across England, from April 1st to June 5th. Note: all data was missing for the 29th of March and the 24th of May. Several hospitals reported values consistent with 100% occupancy (black).

#### Sustainability and Transformation Partnerships (STP) level Bed Occupancy

Across the 42 STPs (aggregates of trusts), there were 20 STP-days (0·7% of total days at risk) where general and acute bed occupancy exceeded 85% occupancy of surge capacity. The highest any STP reached for G&A bed occupancy was 92·7%. For beds compatible with mechanical ventilation, there were 35 STP-days (1·2%) where occupancy exceeded 85% occupancy of surge capacity, 11 STP-days (0·4%) in excess of 92% occupancy, and 4 STP-days (0·1%) at full occupancy (all of which were for STPs outside London: 1) Somerset, 2) Suffolk and North East Essex, and 3) Shropshire, Telford and Wrekin; SFigure 8). Figure 5 illustrates the number of STPs operating at each distinct occupancy threshold as a proportion of baseline and actual surge capacity. The full time-lapse for occupancy over the period of interest can be found in the supplementary material. A similar pattern was seen in the context of critical care / HDU & ITU beds across the STPs (SFigure 5 & SFigure 6).

**Figure 5:**
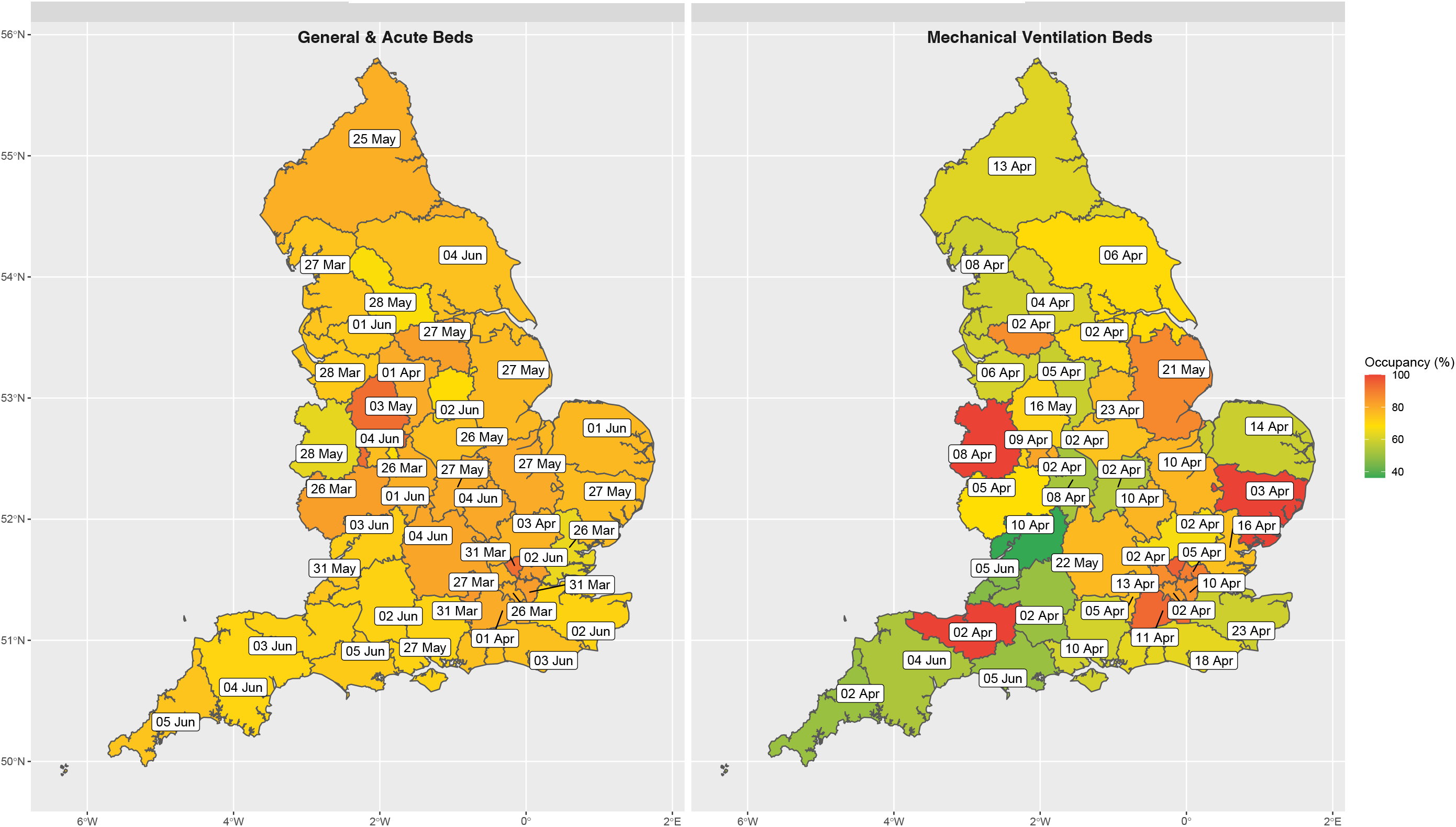
Peak Sustainability and Transformation Partnership (STP) Bed Occupancy Across England. Legend: Figure 5 illustrates the date on which general and acute bed occupancy (Left) and mechanical ventilator beds (Right) peaked, based on surge capacities at the Sustainability and Transformation Partnership (STP) level, across England. The geo-temporal pattern of peak occupancy clearly demonstrates that there was always residual G&A capacity at the STP level, and that all regions across England experienced similar levels of saturation. However, saturation of mechanical ventilator beds differed substantially by location.

### Field (Nightingale) Hospital Occupancy

Of the reported bed capacity achievable through opening the Nightingale hospitals, at peak occupancy only 1·23% of the theoretical maximum were being utilized (Table 2). This equates to 618 bed days for patients with COVID-19 requiring mechanical ventilation, and 1483 bed days for all other types of intervention for patients with COVID-19 (i.e. oxygenation, non-invasive respiratory support, non-respiratory organ support, etc.).

### Independent Sector Care Providers

Variations in reporting meant that the number of providers reporting each day varied, median 181 providers (range: 172 to 187). At peak occupancy, no more than 134 independent sector beds were occupied with patients who were confirmed COVID-19 positive. With regards to patients without COVID-19, at peak occupancy there were 1350 people in independent sector beds, representing a peak saturation of 18·7% (based on the total number of beds reported during contractual negotiations). In summary, there were 3360 bed days for patients with confirmed COVID-19 accommodated by the independent sector (86 mechanical ventilator bed days, 104 non-invasive ventilation bed days, and 3170 other bed days), and 53937 bed days for patients without COVID-19 (2771 mechanical ventilator bed days, 2046 non-invasive ventilation bed days, and 49120 other bed days), between 2nd April and 5th June across England.

## Discussion

This national study of hospital-level bed occupancy provides unique and timely insight into the impact of COVID-19 on bed-specific resource utilization across an entire country. Our analysis suggests that the response of the NHS and British government to COVID-19 was sufficient to alleviate early concerns regarding the number of mechanical ventilators and critical care beds at a national level, however local variation in demand still meant that many trusts reached 100% capacity for both. Moreover, examining occupancy in the context of different organizational units (i.e. trust-level versus STP-level), suggests that the higher-order networks (i.e. STPs) were not efficiently utilized to off-load disproportionately impacted trusts, as it was theoretically possible to have 95·1% fewer trust days at 100% mechanical ventilator bed capacity assuming load was equally distributed. On the other hand, despite a reduction in overall capacity, G&A bed occupancy-levels relatively infrequently reached ‘unsafe’ levels, even at the individual hospital-level. This in part may explain why the field hospitals and independent sector care provider beds were never substantially utilized. Only a very small fraction of the theoretical maximum field hospital bed capacity was operationalized (1·23%). Similarly, despite signing a 14 week block contract with all of the major independent sector care providers valued at £235 million,^28^ these beds too remained largely unoccupied, with less than 24% of the theoretical maximum beds days for established ventilators (i.e. not including the 1012 theatre-specific mechanical ventilators) having been used.

### Context

Initial estimates suggested that an additional 30,000 mechanical ventilators would be necessary to accommodate the impact of the COVID-19 pandemic. These estimates were later updated to just 18,000 mechanical ventilators, from an estimated baseline of 8,000 across the UK.^29^ It is difficult to determine the accuracy of these projections, as they were made in the absence of the impact of non-pharmacological interventions. However, the results of our study suggests that, at the population level, UK-based models of ventilator and bed resource utilization which integrated the impact of non-pharmacological interventions were actually remarkably accurate.^16,30^ Arguably the most influential modelling study was that of the Imperial MRC group, wherein the authors clearly illustrate that with full ‘lockdown’ (i.e. the suite of non-pharmacological interventions that were eventually instituted), that critical care bed capacity would not be overwhelmed.^16^ The nuance that this modelling study lacked was that it failed to explicitly incorporate the impact of unequal distribution of burden, which manifested in our data as specific hospitals and trusts reaching full occupancy, despite the fact that at the national-level there was a substantial number of unoccupied beds.

This retrospective analysis also highlights some of the early incorrect assumptions made about the UK’s baseline resource availability. For example, in contrast to ministerial statements suggesting that there were approximately 8,000 ventilators in the UK prior the pandemic,^29^ our results identified only 4123 operational beds compatible with mechanical ventilation on the first day of reporting in England. Even after acknowledging that our value does not account for the devolved nations (Wales, Scotland, and Northern Ireland), it is unlikely that the initial figures reported by members of parliament truly reflected operational capacity, as that would suggest only 50% of such equipment was in England, despite it representing 84% of the UK population. Interestingly, the absolute increase in ventilator numbers due to the government incentives (e.g. the UK’s Industrial Ventilator Challenge) is much more similar to our reported results.

### Strengths and Limitations

There are several strengths to this study. For example, the use of an administrative (i.e. ‘SitRep’) data that is a statutory collection by NHS England, via a well-established reporting mechanism that has been exploited for research,^31^ confers robustness to the data. One example of how this robustness manifested is, unlike other attempts to collect data at a national level to inform the COVID-19 response plan in the UK,^32^ the degree of missingness in the data utilized in this study was minimal (see supplementary material). Moreover, in light of the unique access to the raw ‘SitRep’ data, we have been able to present our results not only at the trust-level, to which previous endeavors have been limited,^33^ but rather have been able to present information at a much more granular layer (i.e. hospital/site-level) thus providing a much richer understanding of resource utilization that is less prone to the diluent effects of higher level geographies. Finally, a further strength of this study is the relative simplicity of the analysis; there are no complex statistical methods utilized as the descriptive summaries presented are sufficient to describe the experiences of nationalized (single-payer) health system in a high-income economy during the first wave of the COVID-19 pandemic.

Notably though, there are also several limitations to the dataset and our analysis. Firstly, the changes introduced in ‘SitRep’ data collection half-way through the reporting period limited our ability to investigate critical care bed occupancy which was the third bed-specific potential concern identified by forecasting experts. The hospital-level results should also be interpreted in the context of the fact that it is an incomplete representation of the core trust-level information, and thus may not truly reflect the exact position of each organization; for example, the trust corresponding to the single site that achieved 100% G&A occupancy was never itself at 100% occupancy. On a related note, the core weakness of the ‘SitRep’ data is that it presents data as a daily snapshot (at 08:00/8am), and therefore is unable to capture the nuances of the hospital throughput; in essence, both under and over-reporting of occupancy is possible using this method. As such, any marginal results where hospitals are only just over one of the ‘safe occupancy-level’ thresholds should be interpreted with caution as they could represent reporting artefacts. Moreover, the use of the occupancy thresholds reflects a limitation of our analysis, in that a proxy for adverse outcomes had to be utilized given that the necessary information was not readily available to directly explore the relationship between occupancy and patient-level outcome. Finally, the results of this study may not be generalizable to other countries given that it is specific to the UK National Health System infrastructure.

### Implications for Policy Makers, and Clinicians

This study illustrates the potential for near real-time results reporting by which to determine the need for and effectiveness of government policies introduced to address resource utilization-specific issues as a consequence of the COVID-19 pandemic. For example, due to an unequal distribution of the resource utilization burden across England, many trusts spent a significant period of time operating above ‘safe-occupancy’ thresholds, despite the fact that in the vast majority of circumstances there was relief capacity in geographically co-located trusts (i.e. at the STP-level). For illustrative purposes, if load were perfectly re-distributed across all STPs, instead of 23 trusts reaching saturation of their mechanical ventilation beds only 4 would have, and the number of trust days at 100% capacity would have been reduced from 81 to 5 (SFigure 9: STP min-max occupancy plots). This reflects a key operational issue for policymakers to address in preparing for a potential second wave, and would have been identifiable if the SitRep data had been utilized for now-casting. Moreover, other policies for which these results may be relevant, include the creation of the Nightingale (field) hospitals, and independent sector network partnership. Our results suggest that the early investment and the creation of an operational field hospital and independent sector network may yield more overtly positive results in the winter, when G&A occupancy-levels regularly exceed 92%,^34^ however, during the first wave of the pandemic they were under-utilized.

## Conclusion

Using administrative data submitted by all secondary care organizations in England, we can conclude that at the national level there was an adequate supply of all bed-types throughout the first wave of the COVID-19 pandemic. However, the burden of need was not equally distributed, and thus in many cases local demand exceeded the supply of beds, especially where it concerned mechanical ventilation. Although several of the policies introduced by the government, both historical (i.e. STPs) and pandemic-specific (e.g. the independent sector block contract), could have potentially addressed this issue, there is evidence that these interventions were not optimally utilized.

## Data Availability

Trust-level data will eventually be published by NHS England as a freely accessible data resource, but outputs have been delayed by the COVID-19 pandemic. For expedited or more granular access, requests will need to be made directly to NHS England (contact via). All code for this study is available on request.

## Acknowledgements

BAM, SJV, and SD are supported by The Alan Turing Institute (EPSRC grant EP/N510129/). JMD is supported by an Independent Fellowship funded by Research England’s Expanding Excellence in England (E3) fund. SJV is supported by the University of Warwick IAA funding. HW is supported by the Feuer International Scholarship in Artificial Intelligence. JMD, NJT, and APM are supported by the NIHR Exeter Clinical Research Facility. We thank NHS Improvement & NHS England for providing access to the SitRep data and Prof. Matt Keeling for establishing data access. Finally, we thank Dr. Bu’Hussain Hayee for his sense-checking of the final draft.

## Contributors

BAM and SJV conceived the study. Analysis was carried out by HW under the supervision of SJV and BAM. BAM drafted the manuscript, with input from HW and SJV. All authors made substantial contributions to the critical revision of the article, and approved the final version for submission. BAM and SJV take responsibility for the integrity of the data and the accuracy of the data analysis.

## Ethics & Governance

Data utilized in this study were made available through an agreement between the University of Warwick and the Scientific Pandemic Influence Group on Modelling (SPI-M), whom were acting on behalf of the British Government. The study was reviewed and approved by the Warwick BSREC (BSREC 120/19-20).

## Guarantor Statement

The corresponding and the senior author had full access to all data and had final responsibility for the decision to submit for publication.

## Patient and Public Involvement

No patients were involved in the design, interpretation of the results, or dissemination of this study.

